# Real-time estimation of *R*_0_ for supporting public-health policies against COVID-19

**DOI:** 10.1101/2020.04.23.20076984

**Authors:** Sebastián Contreras, H. Andrés Villavicencio, David Medina-Ortiz, Claudia P. Saavedra, Álvaro Olivera-Nappa

## Abstract

**Background:** In the absence of a consensus protocol to slow down the current SARS-CoV2 spread, policy makers are in need of real-time indicators to support decisions in public health matters. The Basic Reproduction Number (*R*_0_) represents viral spread rate and can be dramatically modified by the application of effective public control measures. However, current methodologies to calculate *R*_0_ from data remain cumbersome and unusable during an outbreak.

**Objective:** To provide a simple mathematical formulation for obtaining *R*_0_ in Real-Time, and apply it to assess the effectiveness of public-health policies in different iconic countries.

**Study design:** By modifying the equations describing the spread of the virus, we derived a real-time *R*_0_ estimator that can be readily calculated from daily official case reports.

**Results:** We show the application of a time trend analysis of the *R*_0_ estimator to assess the efficacy and promptness of public health measures that impacted on the development of the COVID-19 epidemic in iconic countries.

**Conclusions:** We propose our simple estimator and method as useful tools to follow and assess in real time the effectiveness of public health policies on COVID-19 evolution.

## 1. Background

Several mathematical models have been proposed in recent weeks to fit public databases on the SARS-CoV2 outbreak (Chen et al., 2020; Simha et al., 2020; Calafiore et al., 2020; Yang et al., 2020). Despite their particularities, most of them have the structure of the well-known SIR model proposed by Kermack and McKendrick (1927). Besides the interest in modeling the spread of this virus, there is a need for indexes to evaluate the efforts made to prevent new cases and assess how likely a particular demographic group is to be infected. One of the parameters used for that means is the Basic Reproduction Number *R*_0_, which value represents the number of persons a single infected individual might infect (Perasso, 2018). From its definition, *R*_0_ ≥ 1 indicate the outbreak might have an exponential growth, while *R*_0_ < 1 would account for a disappearing infection. An intuition on how it works is presented on Figure 1.

**Figure 1:**
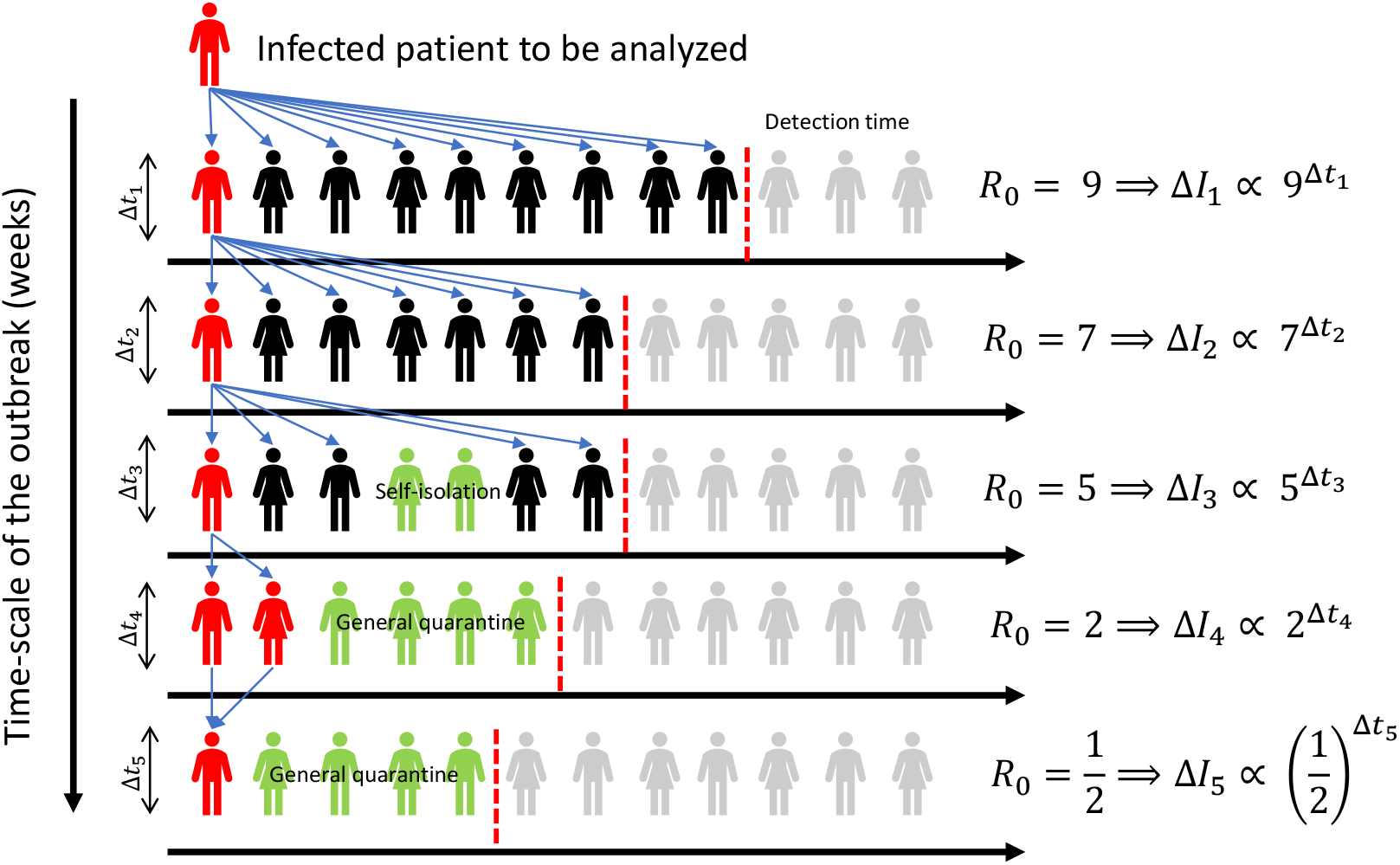
Different stages can be identified during the outbreak of an infection, characterized by the number of people that a single infected individual may infect (*R*_0_). In the figure, a single infected individual (in red) can spread the virus among different individuals (in black), not reaching part of the population (in gray). Some individuals go into isolation (in green), effectively lowering their contagion chance. At the right-hand side of the plot, *R*_0_ represents the number of possible new infections caused by a single patient in each outbreak stage. In the first days of the outbreak, a single individual can infect several people before isolation, but as the amount of cases gets public awareness, health policies restricting movement and self-driven actions may help to control the outbreak, which is effectively captured by a decreasing *R*_0_.

Even though several authors have claimed to have provided guidelines for its calculation (Heesterbeek, 2002; Delamater et al., 2019; Breban et al., 2007; Perasso, 2018), the truth is that it remains uncertain, especially for non-specific public. The formulations presented in literature make the calculation of *R*_0_ nearly impossible for those untrained in mathematical modeling and inverse problems, both because of their complexity and the lack of a general procedure to follow. Then, its objective is not fulfilled, as the different decision-making actors could not use it for evaluating the different actions taken by the different public health plans.

## 2. Objectives

In the present work, we propose a useful and simple methodology to calculate *R*_0_ directly from available epidemiological data in real time during an outbreak. The key feature of this practical methodology is that no specific knowledge in mathematics or scientific computation is needed to generate estimations of this parameter, thus being particularly handy for its use for day-to-day assessment in public health matters. As our methodology does not involve a parameter fitting stage, which would be needed if solving the SIR system numerically to represent continuous trends, we can use it to evaluate the immediate impact of the different actions used to prevent the spread of SARS-CoV2. In a case of study, we assess the effect on *R*_0_ of the different ongoing measures taken by the Chilean government, and we compare their result with the current panorama of different iconic countries.

## 3. Results

Assuming the outbreak follows approximately an SIR model (equations 1-3), susceptible, infected and recovered patient dynamics are represented by:

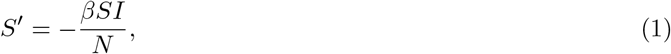

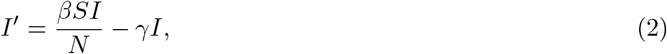

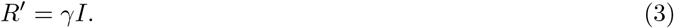

In terms of the parameters of the SIR model, we can calculate *R*_0_ as the ratio between the infection rate *β* and recuperation *γ* (Heesterbeek, 2002):

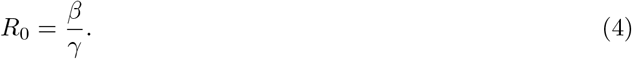

In particular, equations 1 and 2 can be combined applying the chain rule and the derivative of the inverse function theorem, so we can write:

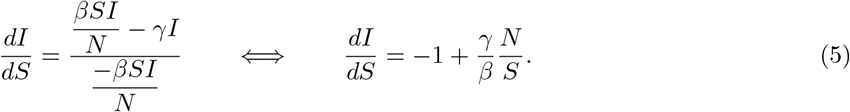

Using the definition of *R*_0_ given by equation 4, 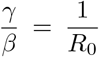. Assuming that all persons are initially susceptible and a low percentage of the population is infected when data is taken, we may safely assume 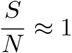. Therefore, equation 5 can be re-written as

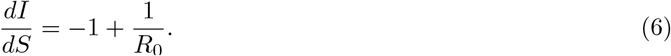

Equation 6 can be discretized in an interval [*t*_*i−*1_, *t*_*i*_] where we can assume that *R*_0_(*t*) = *R*_0_(*t*_*i*_) is constant:

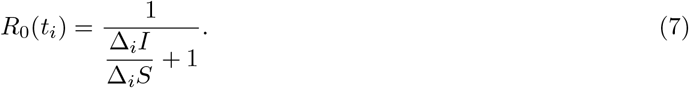

Extending the classical SIR model to consider also deaths, a population balance dictates the discrete differences to follow Δ_*i*_*S* + Δ_*i*_*I* + Δ_*i*_*R* + Δ_*i*_*D* = 0. Then, equation 7 takes its final form.

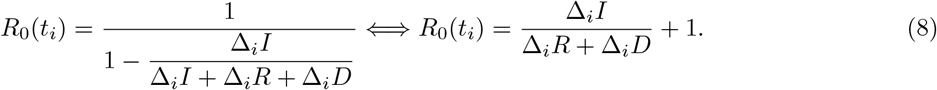

Equation 8 stands in front of other methods because of its simplicity and usability, as there is no need for specific mathematical or scientific computing knowledge for obtaining realistic values of *R*_0_ for a given population during an epidemic or pandemic outbreak. However, due to the nature of its dependence on real-time data, uncertainties on the input values would have a significant effect on the outcome. Since most common uncertainties are related to the time frame between contagion, sampling, detection and report (time misclassification), we suggest applying moving averages to smooth trends and using only official data sources to consistently estimate *R*_0_.

## 4. Discussion

### Effect of quarantine measures on the COVID-19 outbreak in Chile and the current world context

Figure 2 shows the values of *R*_0_ for different countries plotted against time. Countries that have successfully controlled the epidemics currently show *R*_0_ values consistently lower than 1. China, where the COVID-19 pandemics started and is currently under control, shows a slight increase in the last week, which could indicate a new risk factor to be evaluated, such as the presence of a new contagion peak. Other countries currently show lower *R*_0_ values, which may represent better public health managements, with the exception of the current situation in the USA. However, the analysis is much clearer when the *R*_0_ values of all countries are plotted from the day when the first COVID-19 case was reported. In this plot (Figure 2), different control strategies may be straightfully compared. Countries that acted quickly to control viral spread can be recognized by an earlier decrease of their *R*_0_ indexes. The comparative efficacy of control measures can be assessed by the magnitude of the negative slope of the curves. China and Spain reacted at approximately the same time after the first case, but the slope magnitudes of the Chinese plot are noticeably higher and *R*_0_ reaches values below 1 earlier than Spain, and in this last country the contagion rate is not controlled yet. South Korea and Germany controlled the COVID-19 outbreak more or less at the same time, but at much higher rates in the South Korean case, which could reflect different control rate efficiencies of control measures applied in both countries. The differences between Italian and Spanish control policies are also evident, as well as the worsening observed in Italian trends.

**Figure 2:**
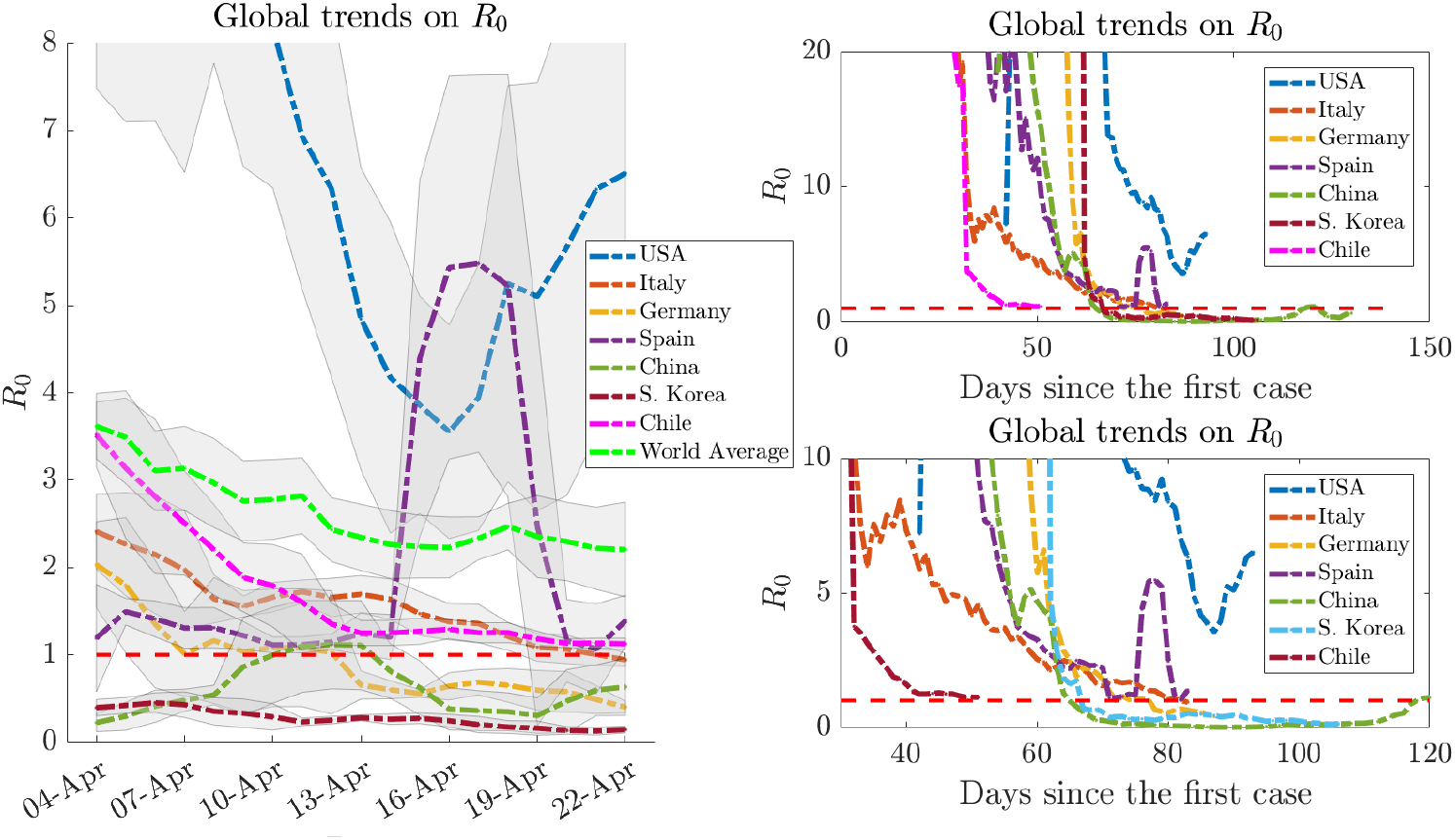
Comparative analysis of the values of *R*_0_ for different countries, using a moving average window of *±* 2 days. The control threshold is represented by a horizontal red dashed line (*R*_0_ = 1). The plot on the left shows daily trends from March 25 to April 22. The rigth-hand charts show the same data plotted from the day of detection of the first case, in two different time frames. Official data from Worldometers.info (16 April, 2020). Files with the data and calculations are available on request.

Using the public database on the SARS-CoV2 outbreak in Chile, provided by the Chilean Health Ministry (MINSAL, 2020a), we studied the numbers up to date (MINSAL, 2020b), and contrasted them to those of countries that have adopted different strategies to control the outbreak. From March 27, the Chilean government declared a partial quarantine for highly affected municipalities in Santiago and other cities (with 42% of the national population and 50% of confirmed cases). Considering an incubation time of 5 days (MINSAL, 2020a), from April 1st on the effects of this quarantine on the SARS-CoV2 spread in Chile can be observed as a decreasing trend in *R*_0_, quickly approaching the control threshold. Up-to-date, both the early onset and high slopes seem to indicate a good effect of the Chilean public-health management policies.

## 5. Conclusions

We have developed a fast and accurate methodology to calculate the Basic Reproduction Number *R*_0_ directly from raw real-time data of an evolving epidemic outbreak. Our results have also shown that this index can be a useful decision parameter to evaluate the impact of public policies in the control of the outbreak of COVID-19. The simplicity of the proposed approach to calculate *R*_0_ (equation 8) remarks its applicability, and our analysis of *R*_0_ trends in different countries during the current SARS-CoV2 outbreak highlights how it can be applied to assess both the speed of reaction and the efficacy of public-health measures. This provides decision-makers with a simple and easily calculable tool to timely understand the impact of their policies. As the proposed equation does not need vast volumes of data, it results particularly handy for its use when data resolution is not high enough to fit continuous models, in the analysis of short-time trends, or to compare different regions in the world or even inside a single country with different time density of data. As varied as the uses for the proposed methodology are the opportunities to improve it. We look forward to seeing how this contribution of a real-time estimator of *R*_0_ would impact the way we analyze the ongoing contingency and how the scientific and decision-making community would adapt it to tailor propagation models and obtain better and timely insights on the application of emergency public-health policies.

## Data Availability

Calculation spreadsheet is available on request

## Conflict of Interest Statement

The authors declare that the research was conducted in the absence of any commercial or financial relationships that could be construed as a potential conflict of interest.

## Author Contributions

Conceptualization, SC, DM-O, HAV; methodology, SC, DM-O, HAV; validation AO-N, SC, CS; investigation, SC, HAV, DM-O; writing, review and editing, SC, DM-O, HAV, AO-N; supervision, AO-N, CS; project administration, AO-N, CS; funding resources, AO-N.

## Acknowledgements

The authors gratefully acknowledge support from the Centre for Biotechnology and Bioengineering - CeBiB (PIA project FB0001, Conicyt, Chile). DM-O gratefully acknowledges Conicyt, Chile, for PhD fellowship 21181435.

## Notes

### Competing Interest Statement

The authors have declared no competing interest.

